# Enhancing Clinical Documentation Workflow with Ambient Artificial Intelligence: Clinician Perspectives on Work Burden, Burnout, and Job Satisfaction

**DOI:** 10.1101/2024.08.12.24311883

**Authors:** Michael Albrecht, Denton Shanks, Tina Shah, Taina Hudson, Jeffrey Thompson, Tanya Filardi, Kelli Wright, Greg Ator, Timothy Ryan Smith

## Abstract

**Objective:** This study assessed the effects of an ambient artificial intelligence (AI) documentation platform on clinicians’ perceptions of documentation workflow.

**Materials and Methods:** A pre- and post-implementation survey evaluated ambulatory clinician perceptions on impact of Abridge, an ambient AI documentation platform. Outcomes included clinical documentation burden, work after-hours, clinician burnout, work satisfaction, and patient access. Data were analyzed using descriptive statistics and proportional odds logistic regression to compare changes for concordant questions across pre- and post-surveys. Covariate analysis examined effect of specialty type and duration of use of the AI tool.

**Results:** Survey response rates were 51.1% (94/181) pre-implementation and 75.9% (101/133) post-implementation. Clinician perception of ease of documentation workflow (OR = 6.91, 95% CI: 3.90 to 12.56, p<0.001) and in completing notes associated with usage of the AI tool (OR = 4.95, 95% CI: 2.87 to 8.69, p<0.001) was significantly improved. The majority of respondents agreed that the AI tool decreased documentation burden, decreased the time spent documenting outside clinical hours, reduced burnout risk, and increased job satisfaction, with 48% agreeing that an additional patient could be seen if needed. Clinician specialty type and number of days using the AI tool did not significantly affect survey responses.

**Discussion:** Clinician experience and efficiency was dramatically improved with use of Abridge across a breadth of specialties.

**Conclusion:** An ambient AI documentation platform had tremendous impact on improving clinician experience within a short time frame. Future studies should utilize validated instruments for clinician efficiency and burnout and compare impact across AI platforms.

## BACKGROUND AND SIGNIFICANCE

Electronic health record (EHR) implementation has resulted in unintended consequences, including increased documentation burden, burnout,^1,2^ and increased stress among clinicians.^3^ Seventy percent of healthcare providers report experiencing stress related to EHR use, with those who report poor/marginal time for documentation having a 2.8 times higher odds of clinician burnout compared with those who had sufficient time.^4^ Ambulatory physicians spend 27% of their clinical day in direct contact with patients while spending nearly double that amount of time on EHR and administrative responsibilities.^5^ Additionally, a study showed primary care physicians need nearly 27 hours in a day to complete recommended guideline-driven care for an average panel of patients, independently increasing workload and stress for clinicians.^6^ Clinicians spend an additional 1 to 2 hours outside of clinical hours performing other administrative work per day.^5^ Working after hours has been associated with increased risk of burnout and lower work-life satisfaction.^2^ There is a pressing need to explore innovative solutions that can alleviate documentation burden on healthcare providers.

The documentation experience varies widely depending on what tool(s) clinicians use, including manual typing, templated text, copy/paste, speech recognition (voice-to-text) technology, and human scribes. In a national cross-sectional study looking at EHR usage, physicians who used predominantly speech recognition technology or transcription spent less time on notes than those who predominantly used templated text, copy/paste, or manual entry.^7^ In addition, a randomized controlled trial in a family practice setting found that in-person human scribes improve time spent charting, perceived chart quality and chart accuracy, and time to note closure.^8^ Human scribes may be helpful for the documentation experience but may be cost-prohibitive for many practices.^9^ Use of these tools may unintendedly add time to a clinician’s workday. For example, speech recognition and manual typing are typically performed after a clinic visit has concluded.

## OBJECTIVE

Ambient artificial intelligence (AI) documentation platforms are emerging technology that use automatic speech recognition and generative AI to summarize a clinical conversation into a structured clinical note. ^10^ As a nascent technology, these platforms have not yet been widely studied in clinical practice. This study examines clinician perception of early use of one such ambient AI documentation tool, Abridge, at an academic medical center, and assesses whether this technology is perceived by clinicians to reduce documentation burden, reduce burnout, improve job satisfaction, and improve note completion time.

## MATERIALS AND METHODS

### Study Design and Participants

We conducted an analysis of survey data administered to physicians and advanced practice practitioners (clinicians) at the University of Kansas Medical Center (KUMC). Clinicians’ perceptions of documentation workflow, risk for burnout, job satisfaction, and timely completion of notes before and after use of Abridge were assessed. The project implementing the AI documentation tool enrolled 181 of 1,255 credentialed medical staff members. Members of physician leadership as well as physician informatics and ambulatory practice committees were prioritized for Abridge (or AI documentation tool) enrollment with word-of-mouth driving subsequent enrollment. Clinicians from 30 medical specialties were included in the study. For analysis, these specialties were grouped into three categories: primary care, medical subspecialty, and surgical subspecialty.

### Ambient AI Platform Description

Abridge is an ambient AI platform that summarizes medical conversations for clinicians and patients across multiple care settings. Clinicians use the Abridge application on their iPhone or Android mobile device to select a patient from their EHR-integrated clinic schedule (Epic). The clinician starts recording with the ability to pause and toggle between different patient encounters as needed. Clinicians then use a web editor to view and edit the AI-drafted note. Evidence that led to the summary can also be called up in a single click that brings up related sections in the transcript and audio to allow for real-time verification and trust in the summary. Once viewing and any editing necessary is complete, dot phrases are used to pull the draft note directly into the clinician’s note template in the EHR.

### Study Approval

The study received a quality improvement designation from the KUMC Institutional Review Board (IRB).

### Survey Instrument

The survey questions were developed by two physician informaticists and a business analyst for quality improvement and operations considerations. The pre-implementation 8-item survey was administered to each clinician just prior to their first use of the AI documentation platform. The overall intent of this survey was to determine the general experience of note documentation of each provider before using the new tool. The initial usage date of the technology and, consequently, the date for administering the pre-implementation survey, varied among participants, ranging from April 2023 to February 2024. Another 11-item survey was developed with the intent of examining the clinician experience post-implementation of the ambient AI tool. The purpose of this survey was to assess clinician perception of benefits of the tool, as well as whether use of the tool should be expanded to a larger number of clinicians across the institution. The questions between the pre- and post-implementation surveys were not the same, although several questions were similar. The complete pre- and post-intervention surveys as well as complete survey item response distributions are provided in the online supplementary appendix.

The post-implementation survey was distributed to all clinicians who had completed at least 5 clinical encounter recordings using Abridge. The survey was emailed to clinicians on February 27, 2024, with a follow-up prompt sent on March 1, 2024, and the survey closed on March 2, 2024. Clinicians had been using the AI platform for a median of 92 days prior to taking the post-implementation survey (IQR: 66 to 172 days, min = 16 days, max = 334 days). Clinical documentation workflow tool utilization was assessed via a multiple mark survey item. A 5-point Likert scale ranging from “Strongly Disagree” to “Strongly Agree” was used for survey items that pertained to documentation experience. There was an additional option of “Not relevant to my experience.” Each participant completed all questions for the surveys. The pre-intervention survey was anonymous and, therefore, the total and percentages of each specialty type depict those providers who could have taken the survey, not necessarily those who took the survey. Since the post-intervention survey was not anonymous, the number and percentages of each specialty type depict the values for those who took the survey.

### Statistical Analysis

To compare the two matched pre- and post-survey questions, we used proportional odds logistic regression (POLR). POLR was also used to look at the effect of specialty type and number of days of use of the digital scribe on post-survey responses. We calculated odds ratios (ORs) and 95% confidence intervals (CIs) from the model coefficients. Statistical significance was set at a p-value of < 0.05. Descriptive statistics were also calculated. All statistical analyses were performed in R version 4.3.2^11^ within RStudio.

## RESULTS

Of the initial 181 clinicians who were offered access to the ambient AI documentation platform, 93 completed the pre-implementation survey (51.9% response rate). The post-implementation survey was completed by 101 of the 133 clinicians who were offered the survey (75.9% response rate). Table 1 depicts the number and percentages of specialty types represented in the surveys.

**Table 1.** Provider Specialties.

| Specialty Type | Pre-Intervention N (%) | Post-Intervention N (%) |
| --- | --- | --- |
| Primary care | 39 (22%) | 34 (34%) |
| Medical subspecialty | 92 (51%) | 41 (41%) |
| Surgical subspecialty | 46 (25%) | 26 (26%) |
| Other | 4 (2%) | 0 (0%) |
| Total | 181 (100%) | 101 (100%) |

Before using the AI platform, 82.8% of respondents agreed or strongly agreed that they regularly spent time documenting outside of clinical hours and 75.2% agreed or strongly agreed that they were at risk for burnout due to documentation. Over sixty percent of respondents cited speech recognition, manual typing, and templates and dot phrases as the predominant tools used for clinical documentation (Figure 1).

**Figure 1.**
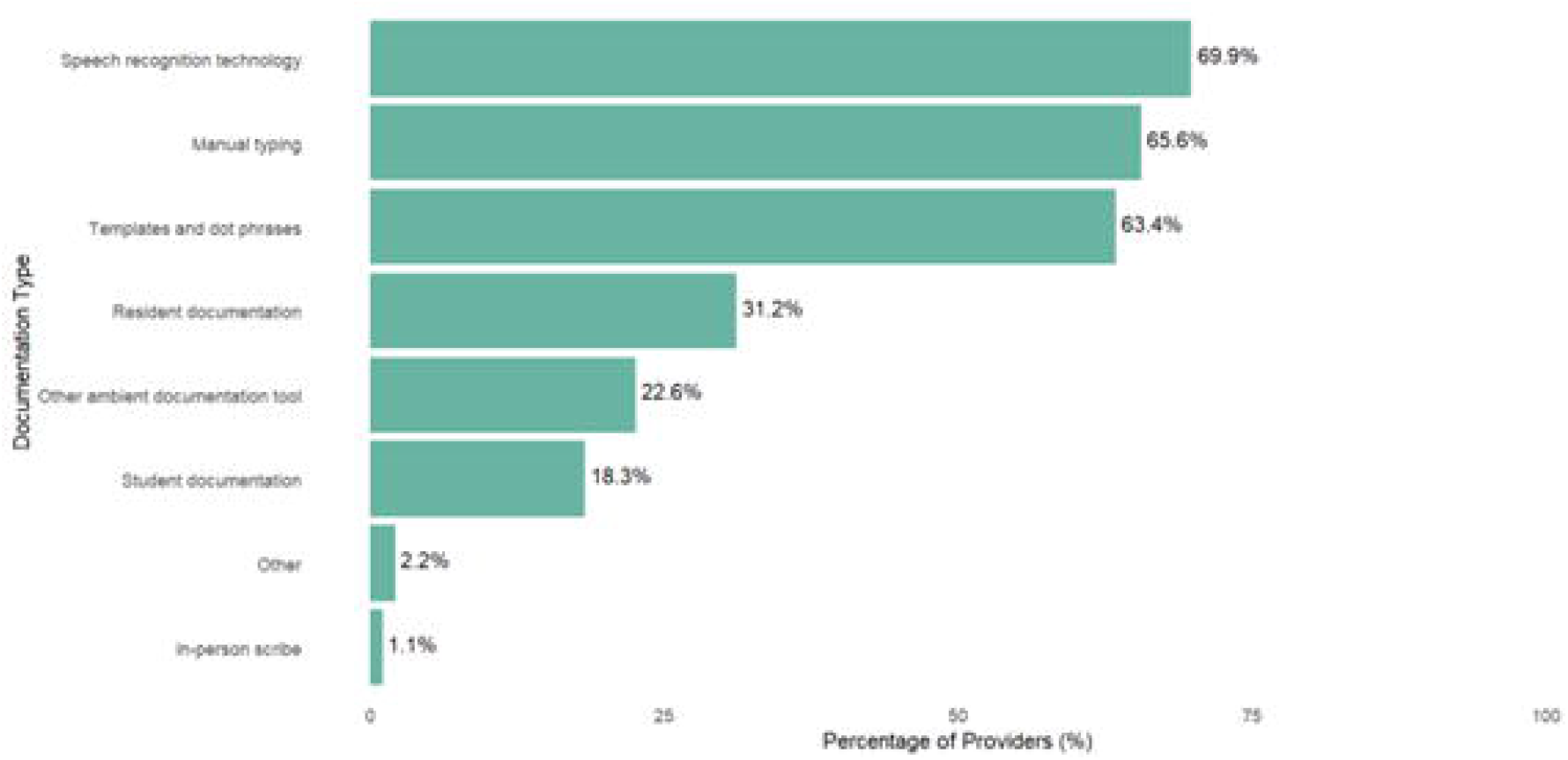
Percentage of clinicians utilizing various documentation workflow tools at baseline. **Alt text:** Horizontal bar graph depicting the percentage of clinicians that used various documentation workflow tools at baseline. The documentation tools used from most frequent to least frequent were speech recognition technology, manual typing, templates and dot phrases, resident documentation, other ambient documentation tool, student documentation, other, and in-person scribe.

Concordant questions between pre- and post-implementation surveys were analyzed collectively to assess the impact of the ambient AI tool on clinical workflow. There was a 6.91 times higher odds of clinicians reporting a higher level of agreement that they find the documentation workflow easy in the post-intervention group compared to the pre-intervention group (CI: 3.90 to 12.56 p< 0.001). There was a 4.95 times increased odds of clinicians reporting a higher level of agreement with the statement that they could complete the note before the next patient visit (CI: 2.87 to 8.69, p<0.001). Table 2 displays a comparison of the concordant questions between the pre- and post-implementation surveys.

**Table 2.** Proportional odds logistic regression analysis comparing concordant pre- and post-intervention survey items.

| Pre-Intervention Survey Item | Post-Intervention Survey Item | Odds Ratio (95% CI*) | P-value |
| --- | --- | --- | --- |
| I find my current documentation workflow easy to use. | Abridge has made my current documentation workflow easy to use for patient visits. | 6.91 (3.90-12.56) | <0.001 |
| I usually complete the note before the next patient visit. | With Abridge, I could complete the note before the next patient visit. | 4.95 (2.87-8.69) | <0.001 |

\*CI = Confidence Interval

Figure 2 displays a descriptive summary of responses from the post-survey. In the post-implementation survey, respondents agreed or strongly agreed 81% of the time that the ambient AI platform had made their current documentation workflow easy to use, enabled completing the note prior to the next visit (43%), improved clinician perception of patient care through decreased documentation burden (77%), decreased the time spent documenting outside clinical hours (73%), reduced the risk for burnout due to documentation (67%), and increased satisfaction at work (64%). Forty-eight percent of respondents agreed or strongly agreed that at least one more patient encounter could be added to clinic sessions if urgently needed. POLR analysis on post-implementation survey questions revealed that post-implementation survey responses did not differ significantly by specialty type (primary care, medical subspecialty, or surgical subspecialty) or number of days of use of the AI platform.

**Figure 2.**
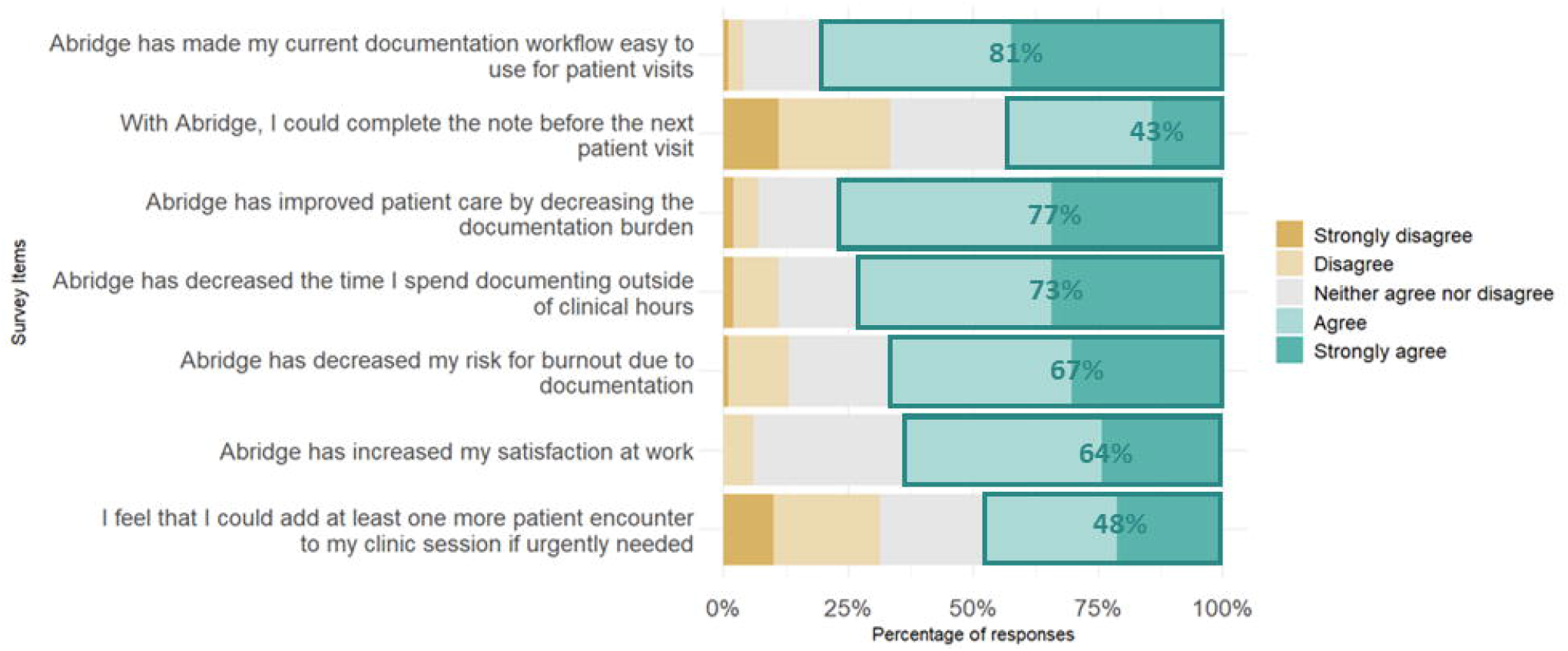
Post-intervention survey response percentages. **Alt text:** Stacked horizontal bar graph depicting percentages of responses to the Likert items on the post-intervention survey.

## DISCUSSION

To our knowledge, this is the first multi-specialty study evaluating clinician perspectives on use of a fully ambient AI platform for clinical documentation. Use of the platform was associated with 7 times greater odds of clinicians reporting a higher level of agreement that their documentation workflow is easy and 5 times greater odds of clinicians reporting a higher level of agreement that they can complete documentation before the next patient encounter. In addition, the majority of clinicians surveyed stated that using the AI platform increased satisfaction at work, decreased documentation burden, decreased time spent documenting outside of clinical hours, and decreased risk for burnout. There was no evidence of a difference across responses when stratified by specialty type or length of use of the AI tool. Furthermore, the positive effects in this study were noted despite approximately 70% of providers already using speech recognition technology at our institution at baseline.

There are several reasons why the ambient AI tool could have been perceived by clinicians to be beneficial in our study. First, the lack of multitasking (as may be required when documenting or using speech recognition while with the patient) and reduced need to recall encounter details by memory may have allowed clinicians to provide more dedicated attention to their patients. Providers may have sensed a greater amount of face-to-face engagement with patients, as ambient AI platforms allow for less interaction with a computer during ambulatory encounters. Second, features inherent to the tool itself such as deep integration of ambient AI into the EHR-based workflow could have also contributed to the noted improvements in workflow ease and note completion efficiency. Third, the reduction in after-hours documentation time attributed to ambient AI usage may have alleviated the strain on clinicians’ work-life balance, potentially reducing burnout risk. The perceived increase in job satisfaction could stem from the sense of support and assistance provided by AI tools in the often-demanding environment of clinical practice. Overall, the multifaceted benefits reported by clinicians highlight the potential of ambient AI documentation platforms to address longstanding challenges in healthcare documentation and improve clinician well-being.

The survey results on the impact of ambient AI documentation tools reveal a divide among providers regarding their ability to see more patients during a clinic session. Approximately half of the respondents felt confident they could add at least one more patient without concern. Protection of perceived wellbeing and control of workload may have disincentivized some respondents from affirming the ability to add patients to their schedules. We underscore that the primary goal of implementing such technology should be to alleviate provider burnout and improve work-life balance, rather than solely to increase patient volume. It should also be noted that without a similar baseline pre-implementation survey question, it is not possible from our study to differentiate whether clinicians answered affirmatively that they could add another patient to the clinic schedule was due to the AI tool intervention or some other factor.

There have been few peer-reviewed published studies to date on the effects of ambient AI documentation technology in clinical practice, and they were generally limited to a specific specialty. An observational study of 110 primary care providers using the ambient AI tool (DAX™) reported a trend towards reducing burnout using the Oldenburg Burnout Inventory and a significant reduction in average time spent documenting notes. However, analysis was restricted to only include participants who completed the survey and used DAX greater than 60% of the time for evaluating burnout (28% or 23 participants) and additionally if there was data available on time in notes (22% or 19 participants).^12^ Another study evaluated the impact of DAX™ in a limited pilot of 12 dermatology physicians and physician assistants. While limited by a low survey response rate (60% or 6 clinicians) the authors concluded the majority of respondents were satisfied with documentation turnaround time, and stated the ambient AI tool “significantly improved” the overall quality of experience with patients.^13^ These studies support that ambient AI may be beneficial; however the results may not be directly relatable to our study in that they utilized a different ambient AI vendor with different product features and user experience, and were smaller studies limited to a specific medical specialty.

One single-center cohort study that evaluated the impact of ambient AI platforms on a multispecialty clinician population utilized a mixed AI and human scribe solution (DAX™) and showed mixed positive and negative effects of the technology.^14^ In that study, while there was an association with improved provider engagement, in work relative value units generated by providers, there was also and an increase in the after-hours time spent in the EHR in the intervention group, in contrast to our study which demonstrated decreased time spent in the EHR. Because the AI tool used in the former study required a human reviewer to edit the draft note prior to routing to the provider for final review, there may have been a long enough lag in note turn-a-round time resulting in providers potentially spending after-hours time to finalize notes. Abridge drafted notes in near real-time (median draft note generation time at our institution was 76 seconds in July 2023 and improved to 38 seconds by April 2024), likely allowing providers the ability to complete the note in a timelier fashion, with some completing notes even prior to the next patient visit.

Our study contributes additional evidence that ambient AI technology not only reduces documentation burden and burnout among primary care clinicians but also benefits medical and surgical subspecialists. While our study included a large number of participants, diversity in medical specialty, and over 50% survey response rates, there are some limitations. The surveys used for this study were originally designed for internal institutional business and operations audiences, rather than for formal research. As a result, pre-implementation survey responses were anonymous, and questions asked in the initial and follow-up surveys contained wordage that prohibited direct comparison of all but two survey items, which resulted in analysis to signal trends instead of associations for clinician satisfaction, cognitive load, burnout, and work outside of work. Given the anonymity of the pre-implementation survey, we were unable to account for the dependence in the groups between pre- and post-implementation survey data. This may have led us to underestimate the variability in the data to some degree. However, this should have a negligible effect on the estimated effect of ambient AI use on documentation experience, which showed a strong association. Ideally, the survey items should have undergone content validation prior to deployment. Future iterations of similar research could benefit from standardized questions across surveys with validated measures for satisfaction, cognitive load, and burnout, use of EHR-based efficiency data, and a control group to better infer causality from the observed changes. In addition, the early adopter cohort of the AI ambient tool may have differed in clinical workload, comfort with technology, burnout, and documentation burden compared to non-participating clinicians. Results may not generalize to clinicians less interested in AI or healthcare information technology. Finally, as this study was limited to a single academic medical center, the results may not be generalizable to other institutions or practice settings.

The present study focused on the subjective perceptions of clinicians on the use of ambient AI documentation platforms in clinical practice. Future research could report on more objective measures related to AI documentation technology, such as exploring time spent on documentation, time to note closure, note quality, number of patients seen, time spent in the EHR outside of work, and effects on revenue generation. This study also focused on short-term use of these tools (less than 1 year for all participants). Longer-term studies of this technology are needed to understand the degree to which there is sustained impact. Studies comparing impact across available and emerging ambient AI tools are needed and can help identify foundational product features needed to maximally support clinicians during their workday. Comparison of ambient AI tools with more traditional technology, such as voice recognition technology and in-person scribes may also be desirable. Furthermore, because this technology is evolving rapidly, often over weeks to months, in quality of note generation and new product features that for example customize clinical documentation to the specific needs of each specialty, research will need to be conducted at a more frequent cadence to properly evaluate impact on clinicians, patients and the greater healthcare ecosystem.

## CONCLUSION

Abridge was associated with a dramatic improvement in healthcare providers’ perceptions of their clinical documentation efficiency, work satisfaction, and risk of burnout across a breadth of specialties. While ambient AI holds early promise to return time to clinicians and curb the burnout epidemic, caution should be raised in tying use of this technology to productivity expectations for an already overworked population. Further studies using validated measures for workflow efficiency, burnout and other aspects of clinician experience are needed, and should be utilized to compare impact across available ambient AI platforms.

## Supporting information

Abridge Pre Survey

Abridge Post Survey

## Data Availability

All data produced in the present study that are not included in the manuscript are available upon reasonable request to the authors.

## ACKNOWLEDGEMENTS

We would like to thank Dr. Daniel Parente for review of the manuscript. We also thank Stephanie Brown and Victoria Bezzole for assistance with survey distribution.

