## Supplementary material for "Enhancing Clinical Documentation Workflow with Ambient Artificial Intelligence: Clinician Perspectives on Work Burden, Burnout, and Job Satisfaction": Abridge Pre Survey

A A A

THE UNIVERSITY OF KANSAS HEALTH SYSTEM

### Abridge Pre-Survey

**Please complete this brief survey about your current documentation experience. It should take less than 5 minutes to complete.**

**Thank you!**

**What is your current documentation workflow in O2?**

**(Please select all that apply.)**

* must provide value

- ☐ Student documentation
- ☐ Resident documentation
- ☐ Manual typing
- ☐ Templates & dotphrases
- ☐ Dragon dictation to desktop
- ☐ Dragon dictation to Haiku/Canto
- ☐ In-person scribe
- ☐ AI & virtual scribe ambient documentation
- ☐ AI only ambient documentation
- ☐ Other

**I find my current documentation workflow easy to use for patient visits.**

**(Please select the best response.)**

- ○ Strongly agree
- ○ Agree
- ○ Neither agree nor disagree

* must provide value

- ◯ Disagree
- ◯ Strongly disagree
- ◯ Not relevant to my experience

**I usually complete the note before the next patient visit.**
**(Please select the best response.)**

* must provide value

- ◯ Strongly agree
- ◯ Agree
- ◯ Neither agree nor disagree
- ◯ Disagree
- ◯ Strongly disagree
- ◯ Not relevant to my experience

**Documenting in O2 negatively impacts my patient care.**
**(Please select the best response.)**

* must provide value

- ◯ Strongly agree
- ◯ Agree
- ◯ Neither agree nor disagree
- ◯ Disagree
- ◯ Strongly disagree
- ◯ Not relevant to my experience

**I regularly spend time documenting outside of clinical hours (after hours/pajama time) because there's not enough time during clinical hours.**
**(Please select the best response.)**

* must provide value

- ◯ Strongly agree
- ◯ Agree
- ◯ Neither agree nor disagree
- ◯ Disagree
- ◯ Strongly disagree
- ◯ Not relevant to my experience

**Documenting outside of clinical hours bothers me.**
**(Please select the best response.)**

- ◯ Strongly agree

* must provide value

- Agree
- Neither agree nor disagree
- Disagree
- Strongly disagree
- Not relevant to my experience

**Generally, documentation causes me stress. (Please select the best response.)**

* must provide value

- Strongly agree
- Agree
- Neither agree nor disagree
- Disagree
- Strongly disagree
- Not relevant to my experience

**I am at risk for burnout due to documentation.**

**(Please select the best response.)**

* must provide value

- Strongly agree
- Agree
- Neither agree nor disagree
- Disagree
- Strongly disagree
- Not relevant to my experience

**Is there anything else about your documentation experience you'd like to share at this time?**

Submit

Powered by REDCap
