## Supplementary material for "Enhancing Clinical Documentation Workflow with Ambient Artificial Intelligence: Clinician Perspectives on Work Burden, Burnout, and Job Satisfaction": Abridge Post Survey

A A A

**Please complete the survey below.**

Thank you!

**Please provide your name.**

* must provide value

**What is your current documentation workflow in O2? *(Please select all that apply.)***

* must provide value

- ☐ Student Documentation
- ☐ Resident Documentation
- ☐ Manual Typing
- ☐ Templates & Dotphrases
- ☐ Dragon Dictation to Deskop
- ☐ Dragon Dictation to Haiku/Canto
- ☐ In-Person Scribe
- ☐ DAX
- ☐ Abridge
- ☐ Other

**Abridge has made my current documentation workflow easy to use for patient visits. *(Please select one.)***

* must provide value

- ○ Strongly Agree
- ○ Agree
- ○ Neither Agree nor Disagree
- ○ Disagree
- ○ Strongly Disagree
- ○ Not Relevant to My Experience

**With Abridge, I could complete the note before the next patient visit. *(Please select one.)***

* must provide value

- Strongly Agree
- Agree
- Neither Agree nor Disagree
- Disagree
- Strongly Disagree
- Not Relevant to My Experience

**Abridge has improved my patient care by decreasing the documentation burden. *(Please select one.)***

* must provide value

- Strongly Agree
- Agree
- Neither Agree nor Disagree
- Disagree
- Strongly Disagree
- Not Relevant to My Experience

**Abridge has decreased the time I spend documenting outside of clinical hours (ie, after hours, non-clinic days). *(Please select one.)***

* must provide value

- Strongly Agree
- Agree
- Neither Agree nor Disagree
- Disagree
- Strongly Disagree
- Not Relevant to My Experience

**Abridge has decreased the stress of documentation. *(Please select one.)***

* must provide value

- Strongly Agree
- Agree
- Neither Agree nor Disagree
- Disagree

- ◯ Strongly Disagree
- ◯ Not Relevant to My Experience

**Abridge has decreased my risk for burnout due to documentation. *(Please select one.)***

* must provide value

- ◯ Strongly Agree
- ◯ Agree
- ◯ Neither Agree nor Disagree
- ◯ Disagree
- ◯ Strongly Disagree
- ◯ Not Relevant to My Experience

**Abridge has improved the quality of my documentation. *(Please select one.)***

* must provide value

- ◯ Strongly Agree
- ◯ Agree
- ◯ Neither Agree nor Disagree
- ◯ Disagree
- ◯ Strongly Disagree
- ◯ Not Relevant to My Experience

**Abridge has increased my satisfaction at work. *(Please select one.)***

* must provide value

- ◯ Strongly Agree
- ◯ Agree
- ◯ Neither Agree nor Disagree
- ◯ Disagree
- ◯ Strongly Disagree
- ◯ Not Relevant to My Experience

**How likely are you to recommend Abridge to a friend or colleague? *(Please select one.)***

* must provide value

- ◯ Not Likely
- ◯ Somewhat Likely
- ◯ Very Likely

**I feel that I could add at least one more patient encounter to my clinic session if urgently needed. *(Please select one.)***

* must provide value

- ◯ Strongly Agree
- ◯ Agree
- ◯ Neither Agree nor Disagree
- ◯ Disagree
- ◯ Strongly Disagree
- ◯ Not Relevant to My Experience

**Please share any additional comments on how Abridge has impacted you:**

**Please share any suggestions or idea for how to make Abridge even more valuable to you and/or your colleagues.**

Submit

Powered by REDCap
